# Early economic evaluation of a predictive tool to test CFTR modulator treatment response in people with cystic fibrosis

**DOI:** 10.64898/2026.01.12.26343787

**Authors:** Hanna de Groot, Marlou C Bierlaagh, Cornelis K van der Ent, Renske M T ten Ham

## Abstract

**Background:** Innovative treatments for rare diseases often strain healthcare budgets. Precision medicine can improve care and reduce costs by guiding treatment allocation. One example is the forskolin-induced swelling (FIS) assay, which uses patient-derived organoids to predict-response to Cystic Fibrosis Transmembrane conductance Regulator (CFTR) modulators in people with Cystic Fibrosis (pwCF). However, it remains unclear when and for whom this assay adds most value. Therefore, the impact of assay accuracy and target population on health-benefits and costs needs assessment.

**Research question:** To quantify the impact of sensitivity, specificity and target population on health benefits and costs to inform further development of a predictive assay to guide treatment allocation in pwCF.

**Study design and methods:** An early economic evaluation was conducted using a decision tree and Markov model. Two strategies were compared over a 40-year horizon: (i) treat all pwCF with CFTR modulators and (ii) predict-response using the FIS assay to guide treatment. Scenario analyses varied assay sensitivity, specificity and treatment responsiveness, reflecting subpopulations of pwCF, including rare *CFTR* variants. Outcomes included quality-adjusted life years (QALY), false negative rates and costs. Model inputs were based on literature on pwCF with F508del mutations.

**Results:** The primary analysis yielded a loss of 1.22 QALYs, with €2,16 million cost-savings per patient in the predict-response strategy. Increasing assay sensitivity reduced QALY loss and false negatives while maintaining cost-savings, while specificity had limited effect on outcomes. Lower treatment responsiveness reduced QALY loss and false negatives while maintaining cost-savings.

**Conclusion:** The assay appears most valuable in pwCF with rare *CFTR* variants, where treatment response is uncertain. Improving sensitivity is crucial to prevent QALY loss, especially in high-responder populations like pwCF with F508del. The model provides insights into variables impacting personalized testing and serves as a dynamic dashboard to explore scenarios once clinical data becomes available.

**Key points:**

- This early economic evaluation provides insights in key variables affecting personalized testing to guide CFTR treatment allocation to inform further assay development.
- High assay sensitivity is crucial to prevent QALY loss in high responder CF populations, such as pwCF with F508del.
- The FIS assay appears most value for pwCF with rare CFTR variants and uncertain treatment response.

## 1. Introduction

Drug development for rare diseases faces numerous challenges which complicate patient access. Challenges include small patient populations, uncertain treatment effects and a lack of well-defined clinical endpoints [1, 2]. While initiatives such as the Orphan Drug designation have incentivized drug development, resulting treatments are often expensive and not always cost-effective [3]. To reduce the burden on healthcare budgets and improve access, it is essential to incorporate economic considerations early in the development of new treatments [4, 5]. This includes identifying patients most likely to benefit from treatment, ensuring scarce resources are spent effectively and add value [6].

Precision medicine, in which biomarkers predict individual treatment response, can offer a solution. By guiding treatment allocation, biomarkers can reduce ineffective drug exposure and decrease costs [7]. However, biomarker-driven treatment allocation remains relatively underexplored in rare diseases, particularly from an economic perspective. Economic evaluations can substantiate value assumptions by quantifying health benefits and costs consequences. Early economic evaluations can guide the development of emerging biomarker-based assay by exploring scenarios. This provides insights into required assay specifications and impact on specific target populations [8]. The forskolin-induced swelling (FIS) assay, a biomarker-based assay in intestinal organoids from people with cystic fibrosis (pwCF), offers a case-study for an early economic evaluation of biomarker-based assays [9-11].

Cystic Fibrosis (CF) is a rare disease and caused by mutations in the *CF transmembrane conductance regulator* (*CFTR*) gene affecting over 54,000 people in Europe [12, 13]. CFTR dysfunction causes thick viscous mucus in various organ systems, resulting in lung function decline, pulmonary exacerbation (PEX) events, decreased quality of life and increased mortality [13]. Historically, CF treatment focused on managing symptoms, but CFTR modulators has transformed CF care for those with access [14, 15]. CFTR modulators improve cellular processing and trafficking of variant CFTR proteins to increase CFTR activity, reducing lung function decline, PEX-events and improving long-term outcomes [13]. The most effective therapy, elexacaftor/tezacaftor/ivacaftor (ETI), is approved by the European Medicines Agency (EMA) and U.S. Food and Drug Administration (FDA). Initially, it is indicated for pwCF carrying at least one F508del mutation (~90% of cases) [13]. More recent, the FDA expanded the indication based on *in vitro* data, and the European Commission (EC) recently approved ETI for all pwCF with a non-class I mutation [16]. However, CFTR modulating therapies are costly (often >$255,600 per patient/year) [17]. Given the heterogeneity in treatment response, especially in pwCF with rare *CFTR* variants, a recently developed forskolin-induced swelling (FIS) assay could help predict treatment response and selectively treat those most likely to benefit.

The FIS assay received a letter of support from the EMA, highlighting its significance in CF treatment optimization [18]. However, specifications required for clinical adoption, such as sensitivity, specificity and target population, remain uncertain. Therefore, in this early economic evaluation we aim to quantify the potential health benefits and costs of implementing the FIS assay under varying specifications and in CF subpopulations with distinct responsiveness to CFTR modulating treatment. Two strategies are compared: (1) a *treat-all* strategy and (2) a *predict-response* strategy, treating only expected responders as determined with the FIS assay. The analysis provides insight into the conditions under which the FIS assay is likely to add most value which informs further assay development.

## 2. Methods

By combining a decision tree and Markov-model, an early economic evaluation was conducted comparing two strategies: i) treating all pwCF (*treat-all*) versus ii) selectively treating pwCF with CFTR modulators based on outcomes of an organoid-based FIS assay (*predict-response*). Using the model, health benefits, expressed as Quality Adjusted Life Years (QALYs), costs and the incremental cost-effectiveness ratio (ICER) were calculated and compared between strategies. The ICER represents the additional cost required to gain one full QALY when comparing a novel intervention to its alternative: ICER = (Costs_intervention_–Costs_standard of care_)/(QALY_intervention_–QALY_Standard of care_). The evaluation took a Dutch healthcare perspective, time horizon of 40 years and assumed a budget-neutral introduction of the FIS-assay without sacrificing effect. The model was constructed in R Studio (version 2024.12.0). The script is available from the corresponding author upon reasonable request.

### 2.1 Population and disease severity classification

Benefits and costs were assessed using a hypothetical cohort (N=100) mimicking characteristics of 12-year-old pwCF with a F508del mutation profile [19]. This cohort was chosen based on the indication for CFTR modulator treatment in the Netherlands and availability of clinical data to substantiate the analysis [19, 20]. Disease severity was classified via percent predicted forced expiratory volume in one second (ppFEV1) levels: mild (ppFEV1 > 70%), moderate (ppFEV1 40%-70%) or severe (ppFEV1 < 40%) [19]. The proportion of the cohort starting in mild, moderate and severe was 98%, 1.7% and 0.3% respectively [19].

### 2.2 Model structure

First, a decision tree was constructed to estimate the proportion of the cohort receiving standard of care (SoC) or additional CFTR treatment for both strategies (Figure 1A). Key assumptions for the primary analysis, including FIS assay sensitivity and specificity and CFTR response rate for the F508del population were derived from literature and expert opinion, see Table 1 [10].

**Table 1:**
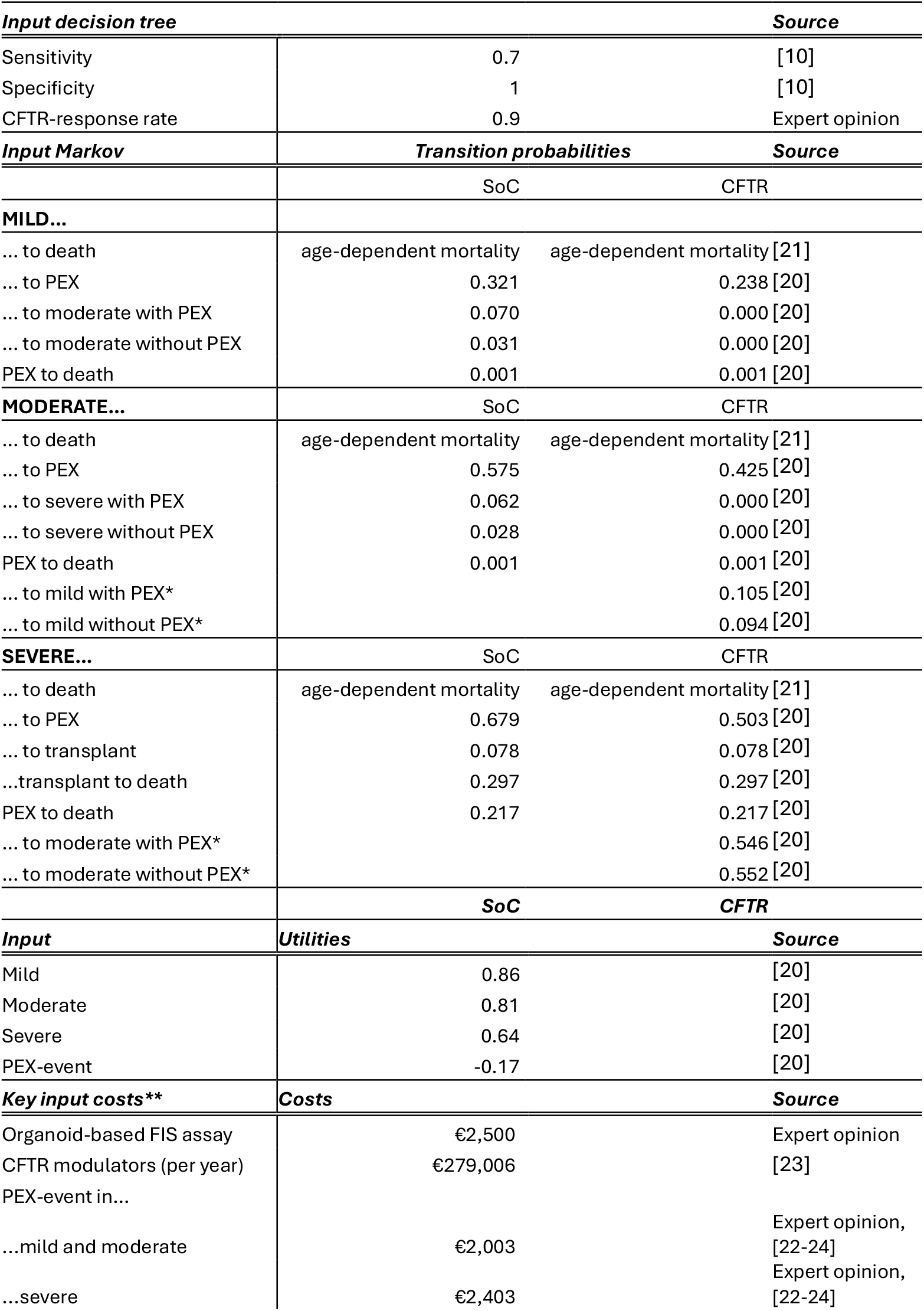
Primary analysis input parameters for decision tree and Markov model. PEX = pulmonary exacerbation. * = With CF transmembrane conductance regulator (CFTR) modulator treatment, patients can only move to a less severe health state in the first cycle. ** Other costs are presented in supplementary Table 1.

**Figure 1:**
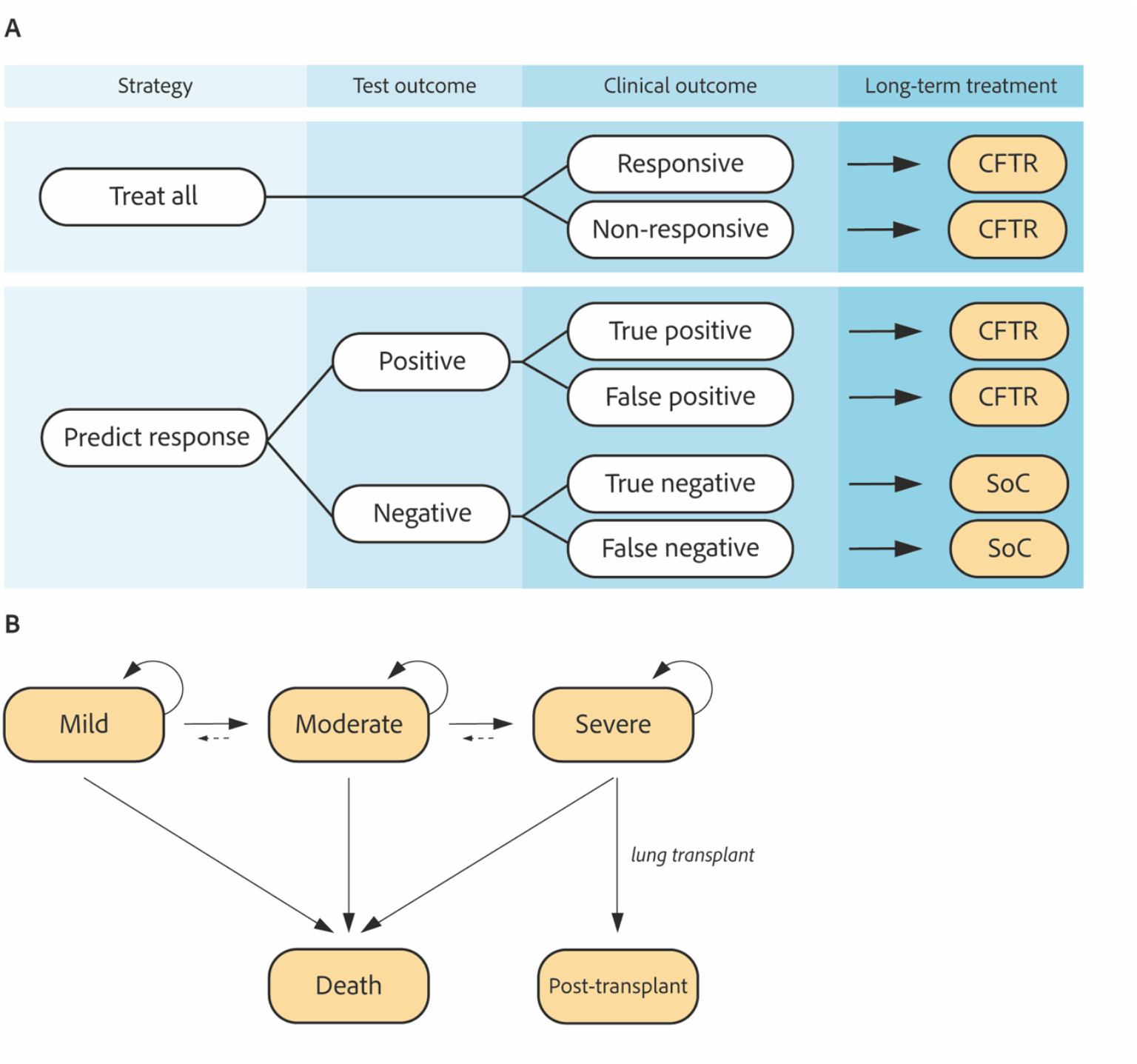
Model design. A) Decision tree design depicting treatment sequence of the treat-all and predict-response strategies. Patients are allocated to either standard of care (SoC) or cystic fibrosis transmembrane conductance regulator (CFTR) modulators. B) Schematic representation of the Markov model structure in which pwCF either receive SoC or CFTR modulators. PwCF enter the model in the mild, moderate or severe health state, whereafter they move through model in yearly cycles. Solid arrows indicate the possible directions of movement. Dotted arrows represent the possibility of moving to a less severe health state which is only possible in the first cycle in the arm receiving CFTR modulators.

Next, to model long-term benefits and costs, a state-transition Markov model with five mutually exclusive health states (mild, moderate, severe, post-transplant and death) was adapted from previous work (Figure 1B) [20]. Based on the decision tree outcome, pwCF either enter the model for SoC or CFTR treatment in the health state matching their disease severity. In each cycle, which represents one year, it was modeled that pwCF remain in their health state or transition to another state in the direction of the arrows (Figure 1B). Within the mild, moderate and severe state, pwCF are at risk of a PEX-event, which increases the probability of disease progression or death [20]. In the severe state, pwCF may undergo a lung transplant, after which they can transition to the post-transplantation or death state. Probabilities of these events are represented by a ‘within cycle’-decision tree [20]. Disease progression after the post-transplantation health state is beyond the scope of this study, meaning that identical to death, pwCF remain in this health state after entering and that no further health effects and costs were considered.

Age and disease-specific mortality rates from 2012 (prior introduction of CFTR modulators) were applied for both treatment groups to exclude CFTR modulator impact on the baseline survival probability [21]. PwCF receiving SoC treatment can only progress to more severe health states (mild → moderate →severe), while pwCF with additional CFTR modulators can improve in the first year (severe → moderate → mild) after which disease progression is halted (dotted arrow, Figure 1B) [20]. CFTR modulators have a relative risk of PEX-events of 0.74 compared to SoC [20]. Transition probabilities are derived from literature (Table 1). Utilities and costs were discounted at 1.5% and 3% respectively according to the Dutch costing guidelines [22].

### 2.3 Input parameters

QALYs were calculated by multiplying the length of survival by utilities. Utilities are standardized health scores ranging between 0 and 1, in which 0 is death and 1 is perfect health [22]. Utilities for mild, moderate and severe states were derived from literature and based on lumacaftor/ivacaftor data (Table 1) [20]. For each PEX-event, a disutility of −0.17 was applied [20]. After lung-transplant, a utility of 0.83 was applied for one year. No further utilities were assigned to the post-transplant state, as studies into the effects of pwCF after lung transplant are lacking and post-transplantation is out of scope of this study.

Yearly health state costs for pwCF in mild, moderate and severe health states were informed by literature [25]. Resource use included clinical visits, imaging, lab tests, lung function tests and medication, and was validated and supplemented via expert opinion (Supplementary Table 1). Additional resource use related to PEX-events, including clinical admission and antibiotics, were informed by expert opinion. Costs for SoC, CFTR modulator treatment (ETI, Trikafta®/Kaftrio®) by Vertex Pharmaceuticals Incorporated, Boston, MA, USA) and PEX-events were calculated using public list prices and reference prices from the Dutch Healthcare Institute and the Dutch Healthcare Authority [22-24]. Lung transplant costs were based on Dutch Diagnosis-Related Group codes [24]. All prices were collected for or converted to 2023 prices (in Euro’s; €) [26].

### 2.4 Assumptions

Several assumptions were made while constructing the model. Assumptions were informed or validated by expert opinion (N=2). First, SoC medication in pwCF receiving CFTR modulators was adjusted based on disease severity. In the mild state, SoC medications were discontinued. In the moderate state, SoC medication usage was reduced by half, while in the severe state, full usage was maintained. Next, during PEX-events, pwCF in mild and moderate disease states received 21 days of home i.v.-antibiotics. In the severe state, 10% underwent an additional 7-day clinical admission. Additionally, it was assumed no PEX-events occurred in the first six months, followed by a maximum of one PEX-event per year thereafter. Last, CFTR treatment effect was assumed to remain constant over time.

### 2.5 Scenario and sensitivity analyses

The impact of sensitivity, specificity and CFTR modulator response rate on total QALYs, costs and rate of false negatives were investigated in scenario analyses (Table 2). CFTR modulator responsiveness served as a proxy for different subpopulations of pwCF, reflecting variation in treatment response due to genetic differences (e.g. rare *CFTR* variants). Three scenarios were formulated in which the (i) sensitivity and (ii) specificity was varied independently from 0.5 to 1.0, and (iii) CFTR modulator responsiveness, which varies between CF subpopulations, was varied from 0.0 to 1.0. Additionally, CFTR modulators are patent protected in most countries until 2037, after which prices are expected to drop due to generic competition [17]. Thus, in scenario (iv), a price reduction of 73% was modelled after 13 years (corresponding to 2037), based on public Argentinian off-patent CFTR modulators prices [17]. Prices were adjusted to Dutch prices using the Purchase Power Parity index [17, 27].

**Table 2:** Overview of scenarios and parameter variations for scenario analyses. CFTR = Cystic Fibrosis Transmembrane conductance Regulator modulators.

| Scenario | Sensitivity | Specificity | CFTR response rate | Price reduction |
| --- | --- | --- | --- | --- |
| 1 variable sensitivity | 0.5 - 1.0 | 1.0 | 0.9 | NA |
| 2 variable specificity | 0.7 | 0.5 - 1.0 | 0.9 | NA |
| 3 variable CFTR response rate | 0.7 | 1 | 0.0 - 1.0 | NA |
| 4 patent expiry | 0.7 | 1 | 0.9 | 73% after 13 years |

Uncertainty around input parameters and assumptions were explored via a deterministic sensitivity analysis (DSA). A DSA explores the impact of individual parameters on the ICER by alternately varying input values between pre-set minimum and maximum values and can be considered parameter-specific best- and worst-case scenarios. Therefore, utilities, costs and sensitivity and specificity of the FIS assay were varied between 80% and 120% from the value used in the primary analysis.

## 3. Results

### 3.1 Primary analysis

In the primary analysis, the *treat-all* strategy yielded 21.88 QALYs and €6.15 million per patient over a time span of 40 years, compared to the *predict-response* strategy, which yielded 20.66 QALYs and €3.99 million per patient. This is a difference of 1.22 QALY lost for €2.16 million saved, with 27% of the tested cohort being false negatives.

### 3.1 Scenario analyses

The impact of varying sensitivity, specificity, and CFTR response rate was assessed across scenarios 1-3 (Table 2, Figure 2). In scenario 1, increasing sensitivity (from 0.5 to 1.0) in the *predict-response* strategy led to more true positives, resulting in higher treatment costs, increased QALYs, and fewer false-negative diagnoses. Thus, increasing sensitivity protects QALY loss. (Figure 2 A-C). In scenario 2, increasing specificity (between 0.5 to 1.0) reduced false positives, ensuring fewer patients were incorrectly identified as responders. However, this had little effect on QALYs and treatment costs (Figure 2 D-F). In scenario 3, increasing the CFTR response rate (from 0% to 100%) raised the number of false positive tests, leading to increased QALYs and costs as the number of individuals benefiting from treatment increased. However, this also increased the false-negative rate (Figure 2 G-I).

**Figure 2:**
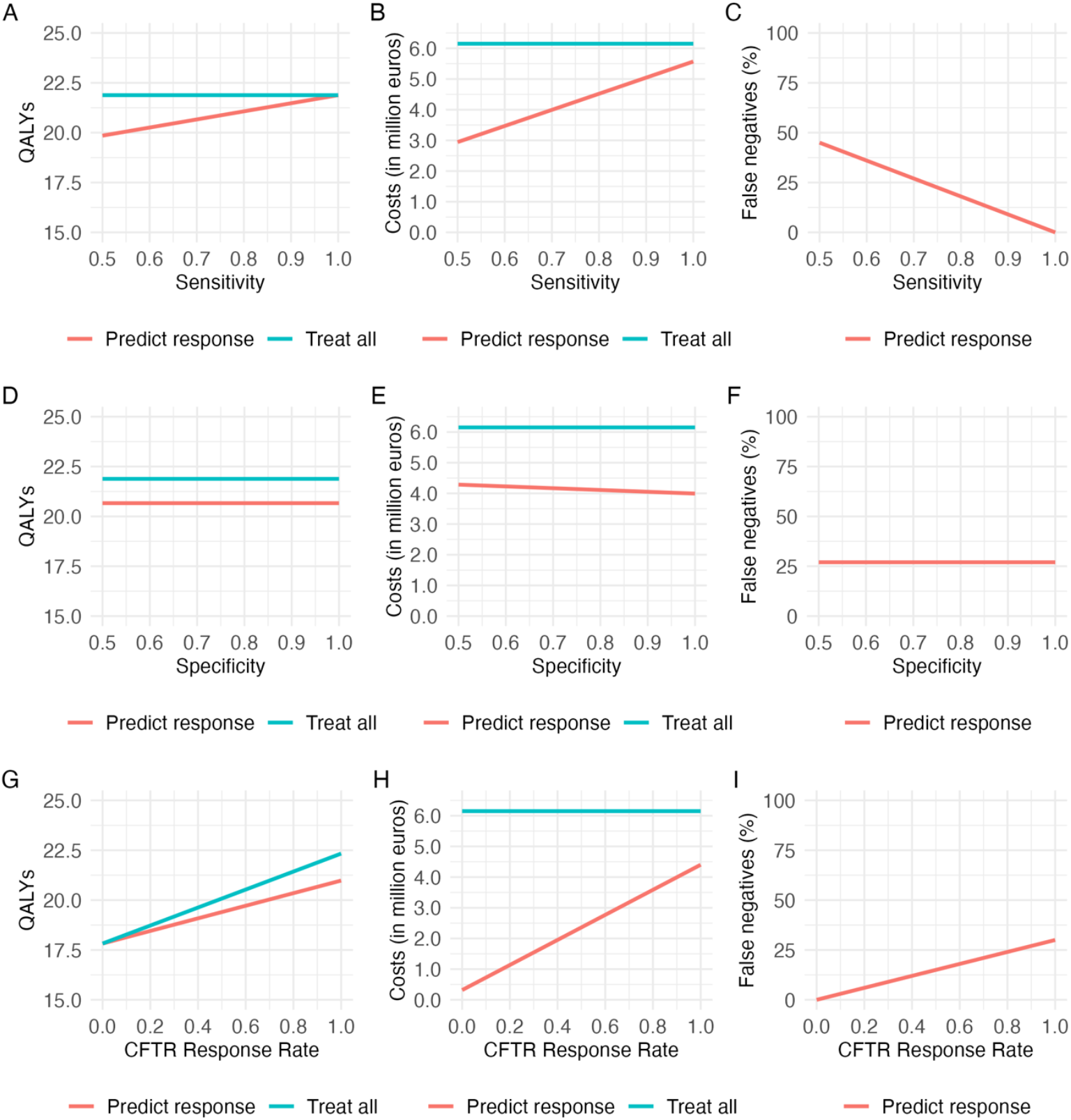
A-I: Outcomes of scenario analyses on the treat-all (blue) and predict-response (red) strategies. QALYs and costs were calculated per patient over a 40-year time horizon. 2A-C: Impact of sensitivity (top), 2D-F: specificity (middle) and 2G-H: CFTR response rate (bottom) on QALYs (left), costs (middle) and false negatives (right). QALY = quality adjusted life years, CFTR = Cystic Fibrosis Transmembrane Conductance Regulator modulators.

Scenario 4 assessed the impact of patent expiry (Table 2, Figure 3). In the *treat-all* strategy, cumulative costs dropped by 2.86 million per patient (€6.15 million to €3.86 million) after patent expiry from year 14 onwards. In the *predict-response* strategy, costs decreased by €1.44 million per patient (€3.99 million to €2.55 million). Costs of the *predict-response* scenario remained lower than the *treat-all* strategy, regardless of patent expiry.

**Figure 3:**
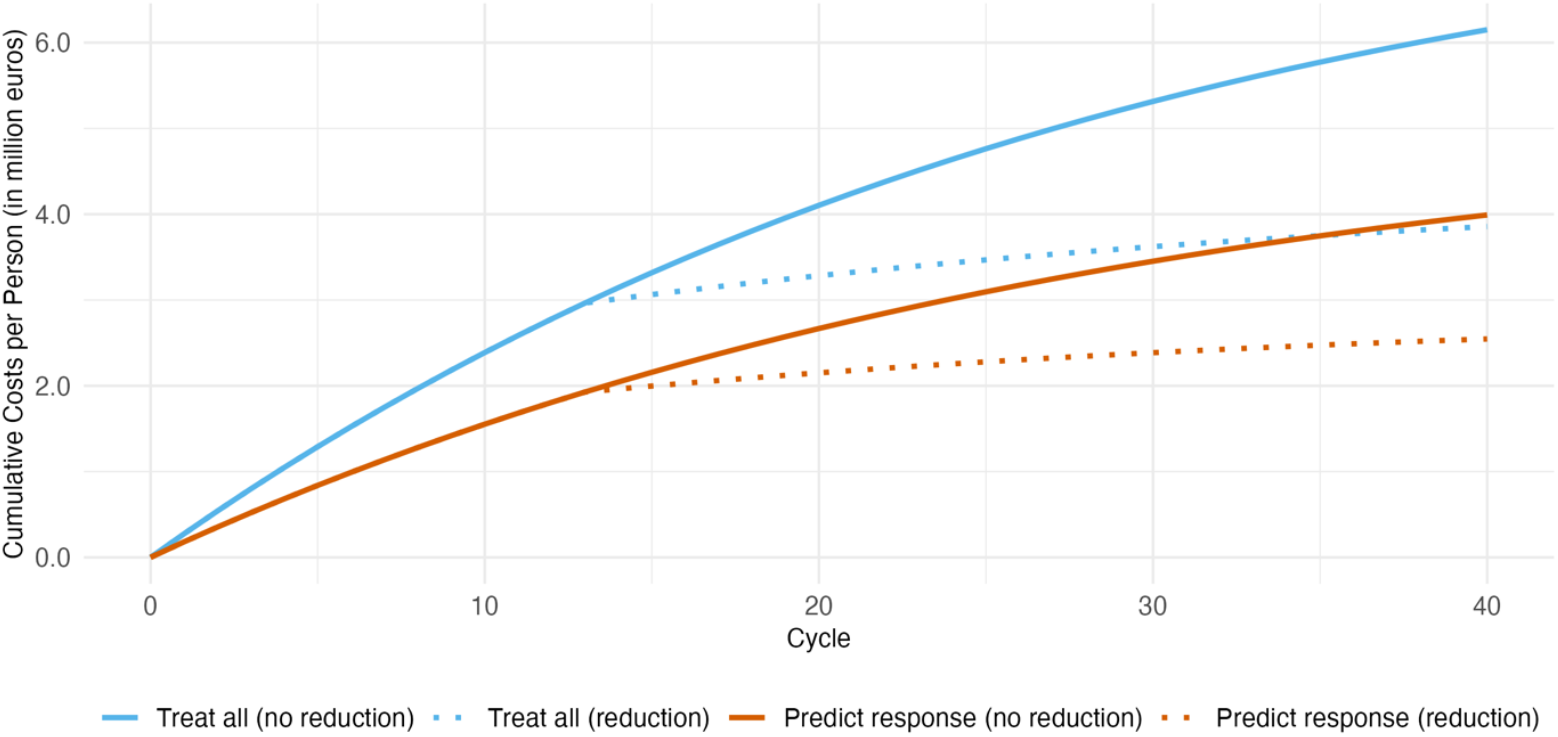
Cumulative costs per patient following patent expiry after 13 years. Costs are shown for the treat-all (blue) and predict-response (red) strategies with (solid line) and without (dotted line) price reduction after 13 years (scenario 4).

### 3.2 Sensitivity analysis

The DSA results display 10 parameters that influence the ICER most (Figure 4). Variation in the utilities in the mild and moderate health state, CFTR modulator response rate, costs of CFTR modulators and sensitivity had most influence. These findings align with scenario analyses outcomes, where changes in sensitivity and CFTR response rate impacted both QALYs and costs.

**Figure 4:**
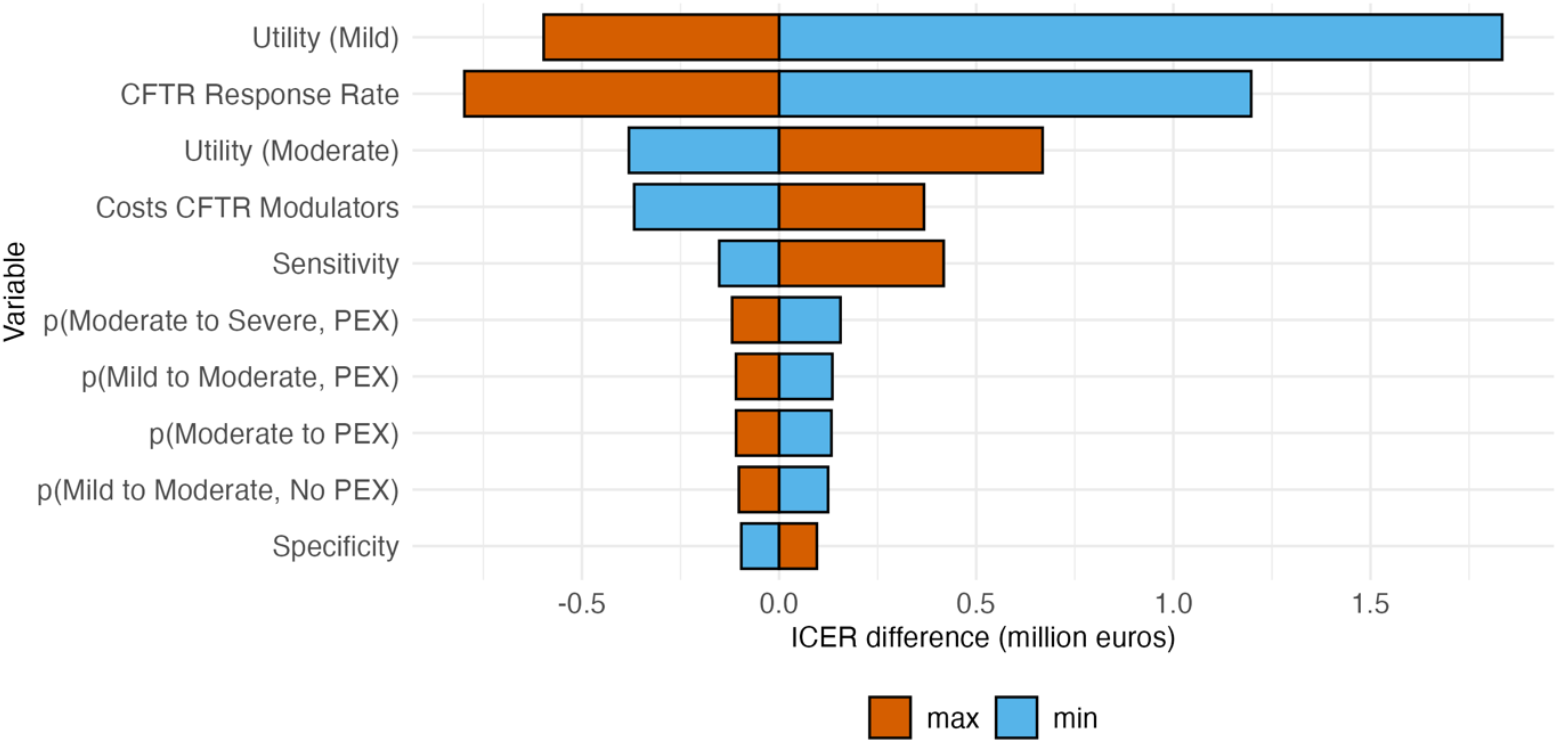
Results of the deterministic sensitivity analysis (DSA) visualized in a tornado plot. The plot visualizes the impact of variation of individual parameters on the incremental cost-eAectiveness ratio by alternately varying input values between 80% (min, blue) and 120% (max, red) of their original value. CFTR = Cystic Fibrosis Transmembrane conductance Regulator, PEX = pulmonary exacerbation, ICER = Incremental Cost-EAectiveness Ratio.

## 4. Discussion

This study presents an early economic evaluation of the FIS assay for guiding CFTR modulator treatment allocation in pwCF. Health benefits and costs consequences of potential assay specifications in pwCF with a F508del mutation were assessed. As this is an early economic evaluation, model outcomes should be interpreted as a tool to support further FIS assay development, rather than as definitive evidence of cost-effectiveness. The primary analysis showed potential cost savings for the *predict-response* strategy of €2.16 million per patient over 40 years, at the loss of 1.22 QALYs in the highly CFTR modulator responsive F508del population. This is reflected in an ICER of €1.77 million per QALY lost. The ICER is most sensitive to changes in utility in mild disease, CFTR response rate of the population and sensitivity of the FIS assay. In our analysis, QALY losses and cost savings were mainly due to 27% of individuals being incorrectly excluded from treatment due to false negative test results. Therefore, improving assay sensitivity is essential for clinical adoption of the FIS assay in high CFTR modulator responder populations, by reducing missed treatment opportunities and preventing QALY loss.

Scenario analyses further explored the effects of varying assay specifications and CFTR modulator responsiveness reflecting characteristics of broader CF subpopulations, where treatment response is uncertain. Among pwCF with *CFTR* mutations not included on the FDA label, response rates to ETI are estimated to be as low as 50% [28]. However, the recent label extension in the EU includes all pwCF with non-class I mutations. In scenarios assuming a 50-60% CFTR modulator response rate, the *predict-response* strategy resulted in significant cost savings compared to the *treat-all* strategy [29]. Equally important, in lower CFTR responsive populations, the number of false negatives also decreases with assay specifications of a sensitivity and specificity of 1.0 and 0.7 respectively. Thus, based our analysis assay implementation subpopulations of pwCF with lower CFTR modulator responsiveness is associated with less QALY loss and greater cost-savings compared to high responder populations.

The predicted QALY loss observed in both the primary and scenario analyses calls for careful ethical deliberation. A critical question remains about the acceptability of excluding CF patients from CFTR treatment based on predictive assay outcomes. Therefore, further assay development must be accompanied with transparent dialogues between clinicians, regulators and patient representatives to discuss acceptable thresholds for false negative rates and cut-off values used to classify patients as responders or non-responders.

Previous research has investigated the cost-effectiveness of a similar predictive tool to guide CFTR allocation decisions in a Canadian F508del population and healthcare setting. Consistent with our primary analysis findings, they found that while CFTR modulators increase patient survival and reduce PEX-events as well as lung transplants, this did not translate into cost savings due to the high medication costs associated with CFTR modulators. In line with our results, an ICER of $0.9 to $1.1 million per QALY lost was reported, which strengthens the validity of our model.

### 4.1 Strengths and limitations

In this study, a decision model was developed which allows exploration and quantification of potential benefits and costs of implementing the organoid-based FIS assay in subpopulations of pwCF. While this is an early economic evaluation, the model provides a dynamic dashboard for assessing the impact of technical and clinical parameters on both patient outcomes and costs. By enabling scenario exploration, it contributes to the ongoing development of the organoid-based FIS assay. Therefore, a strength of our approach is the adaptability of the model, as it can be updated with new clinical evidence to enhance its accuracy. This flexibility allows for future studies to evaluate the effect of different (CFTR) therapies and subpopulations of pwCF more precisely, including those with rare *CFTR* variants. Important new input for the model is currently generated in the CHOICES trial. This study evaluates a new CFTR modulator combination, dirocaftor/posenacaftor/nesolicaftor, designed for both rare *CFTR* variants and the F508del population. In addition, ongoing automation and development are anticipated to enhance the FIS assay’s performance characteristics [30]. Incorporating emerging clinical efficacy and assay performance data in the prediction model could make the model more accurate.

Several limitations of the model must be considered when interpreting the results. First, as this is an early economic evaluation, input parameters are derived from literature, which may not fully represent context-specific characteristics, thereby introducing uncertainty. Transition probabilities and utilities were derived from lumacaftor/ivacaftor trial data instead of ETI, which may cause an underestimation of treatment effect for other (highly effective) modulator therapies. However, external validation with clinicians and CF-registry data of the modeled disease progression in both treatment groups indicate that these are well-suitable to use as an estimate for such effective treatments. In addition, the QALY differences between screening and not screening for drug response, as well as the QALYs observed in the separate CFTR and SoC groups align with existing literature, strengthening the external validation of the model [20, 25]. Second, the model assumes static utility values, yet real-world treatment effects such as side-effects and psychosocial benefits of receiving a life-extending treatment are not captured. As a result, true impact of CFTR modulators on quality of life might be underestimated. Future work should incorporate real-world data and patient-reported outcomes to refine the model’s assumptions. Lastly, since genotype-specific clinical outcomes for rare mutations were unavailable in registry data, clinical data from the F508del population were utilized as a proxy to reflect on possible implications for other, less common variants [13, 19]. Rare mutations show high variability in disease progression, which makes economic modelling challenging. Therefore, the F508del population offers a consistent reference point and provides a structured benchmark. However, differences in transition probabilities could occur in both directions, potentially impacting model accuracy.

### 4.2 Future research

The primary analysis is based on clinical data from the F508del population, where the organoid-prediction model has only demonstrated the ability to distinguish treatment effectiveness at a group level [31]. However, to date, no association has been established between individual response prediction and clinical outcome [32, 33]. The predictive accuracy of the organoid-based FIS assay should therefore be refined for various CFTR modulating therapies in different subpopulations. Second real-world patient data, such as generated in the CHOICES trial, should be incorporated to validate the use of static utilities [34].

### 4.3 Clinical relevance

The recent approval by the EC to extend access to ETI for all pwCF who carrying non-class I mutations marks a significant step forward in expanding treatment availability. However, while this decision allows for market authorization, it does not guarantee reimbursement by national healthcare systems. In the EU, reimbursement decisions are made on a Member State level and are increasingly informed cost-effectiveness analyses. This analysis suggests that using a predictive response model, like the FIS assay, could reduce ineffective treatment and lead to substantial cost savings. Nevertheless, ethical concerns remain regarding the potential exclusion of false negatives.

Ultimately, in further developing the assay it is up to regulatory decision-makers to assess what level of misclassification is acceptable in clinical practice, weighing the balance between cost and patient access to treatment. This threshold may differ from the perspective of healthcare providers, who prioritize individual patient outcomes and may advocate for broader access despite economic constraints. As such, ongoing dialogue between regulatory bodies, clinicians, and patient advocacy groups will be essential in shaping policies around the clinical implementation of predictive testing for pwCF and this model can help the discussion.

## 5. Conclusion

An early economic evaluation assessed the potential health benefits and cost-consequences of various FIS assay specifications in subpopulations of pwCF, identifying conditions under which it is most likely to add value. In subpopulations with high CFTR modulator responsiveness, improving assay sensitivity is critical to prevent QALY loss, despite substantial cost savings. Based on current performance estimates, the FIS assay is most likely to add value in CF subpopulations with lower responsiveness, such as pwCF with rare *CFTR* variants, where both QALY gains and cost savings can be achieved. Ultimately, FIS assay guided treatment allocation can improve patient access to effective treatments, while ensuring efficient use of limited healthcare resources by aligning treatment decisions with both clinical need and economic impact.

## Data Availability

All data produced in the present study are available upon reasonable request to the authors

## Author acknowledgements

HdG, MB, CvdE and RtH designed the study. HdG performed the analysis. HdG and MB took the lead in writing the manuscript. RtH is acting as guarantor. All authors provided critical feedback and helped shape the research, analysis and manuscript.

## Statement and declaration

HdG, MB and RtH declare no conflict of interest. CvdE holds Patent No. 10006904 and receives royalties, with no influence on this study.

## Data availability statement

Data was derived from literature and expert opinion. All input parameters used in this model are included within this paper. R scripts are available from the corresponding author upon reasonable request.

